# The impact of cognitive processes associated with image recognition on visuo-vestibular interaction

**DOI:** 10.64898/2026.04.22.26351361

**Authors:** Pasquale Malara, Andrea Giorgia Tosin, Andrea Castellucci, Salvatore Martellucci, Lucia Belén Musumano, Marco Mandalà

## Abstract

An increasing number of studies highlight the role of saccadic remodulation in compensatory mechanisms following vestibular injury, and the reappearance of SHIMP saccades correlates with symptom improvement measured by the Dizziness Handicap Inventory (DHI). To investigate the influence of attentional processes and working memory on visuo-vestibular interaction, three independent but interrelated experiments were conducted.

In the first two experiments, healthy subjects and patients with unilateral or bilateral vestibular deficits underwent vHIT in SHIMP mode and the Functional Head Impulse Test (fHIT), performed first separately and subsequently simultaneously. Mean latency and clustering of SHIMP saccades, together with Landolt C recognition rates, were analyzed. Differences between separate and combined protocols were assessed, and, in patients, correlated with symptom severity measured by the DHI, to determine whether the near-simultaneous execution of tasks mediated by shared parietal cortical substrates influenced performance.

In the third experiment, vHIT in HIMP mode and fHIT were performed using separate and combined protocols to evaluate whether recognition-related cognitive load affected recovery saccade latency and clustering.

Results suggest that visual recognition modulates visuo-vestibular interaction, supporting integrated dual-task protocols for ecological balance assessment and helping explain clinical discrepancies.

## 1. Introduction

In the past, it was believed that, following a unilateral vestibular deficit, compensatory processes led to a progressive recovery of the gain of the vestibulo-ocular reflex (VOR), thereby restoring image stabilization on the fovea during rotational head movements (1,2,3). More recent evidence instead indicates that functional recovery depends predominantly on compensatory saccadic movements (4,5).

Over time, these saccades undergo adaptive changes characterized by increased clustering and reduced latency, with progressively improved synchrony relative to the residual slow phase of the impaired VOR (6,7).

The introduction of the SHIMP (Suppression Head Impulse Paradigm) modality into the video Head Impulse Test (vHIT) has made it possible to analyze the relationship between the residual slow phase of the VOR and the characteristics of anticompensatory saccades. The early reappearance of these saccades after a vestibular insult is strongly associated with clinical improvement, as demonstrated by Dizziness Handicap Inventory (DHI) scores, even in the absence of a substantial increase in canal gain (8,9,10).

Given the central role of saccades in vestibular compensation processes, it therefore appears relevant to further investigate the cognitive factors that may influence their kinematics.

Numerous studies have demonstrated a relationship between vestibular function and several domains of visuospatial ability, including spatial memory, navigation, mental flexibility, and the mental representation of three-dimensional space (11). Additional evidence has further suggested that the vestibular system exerts a significant influence on attention and executive functions (12,13,14), as well as on several other cognitive domains, as reported by Agrawal et al. (15), Brandt et al. (16), Ekvall Hansson E. & Magnusson M. (17), Guidetti et al. (18), Popp P. et al. (19), Zheng Y., Goddard M., Darlington C.L. & Smith P.F. (20), Risey J. & Briner W. (21), and Taljaard D.S. et al. (22).

The aim of the authors of this article (23) is to demonstrate how the intrinsic informational value of visual inputs significantly modulates the motor commands responsible for visually guided voluntary saccades. In particular, the salience of the stimulus and the degree of cognitive processing associated with and assigned to the saccadic movement influence the kinematic parameters of the saccades themselves, resulting in reduced latency and increased eye movement velocity. This suggests a functional interaction between cognitive processes involved in evaluating stimulus salience and oculomotor control. The study by Minnan Xu-Wilson et al. (24) demonstrates that this modulation also affects visually induced reflex saccades, in addition to voluntary ones. Neural signals reflecting value and action selection in the context of eye movements have been identified in several brain structures: the basal ganglia (25), the posterior cingulate cortex (26), and the amygdala (27). These distributed signals may influence oculomotor output, for example through the basal ganglia, which project directly to the superior colliculus, known for its influence on saccadic movement kinematics. Furthermore, the work of Corbetta et al. (28) highlights that cortical circuits involved in attentional processes largely overlap with regions responsible for oculomotor control; in particular, within the parietal cortex, neural signals related both to attention and working memory and to eye movements coexist. The article emphasizes that some authors, based on this close convergence of cortical areas, go so far as to hypothesize that attentional processes and eye movements are essentially different expressions of how consciousness explores and interprets the surrounding world and are therefore inseparable from one another.

Two additional studies have experimentally investigated the influence of cognitive processes on the kinematics of saccadic movements and on their integration with the vestibular system. In the study by Simon Schwab et al. (29), patients with schizophrenia and healthy control subjects were subjected to visually guided voluntary saccadic tasks with progressively increasing salience load. Participants were required to recognize and compare stimuli presented before and after the saccadic movement, and were free to decide whether to move their heads in order to support the oculomotor action. In the first condition, the stimulus to be compared was a color, whereas in a second condition it was a Landolt optotype, characterized by greater cognitive processing demands and therefore higher salience. In healthy subjects, the increase in salience associated with the oculomotor task resulted in reduced latency (the reported improvement averaged approximately 40 ms) and increased saccadic velocity, confirming the cognitive modulation of eye-movement kinematics. In patients with schizophrenia, this effect was not observed: latency and saccadic velocity remained essentially unchanged despite the increased complexity of the task. Moreover, whereas performance in color recognition was comparable to controls, recognition of the optotype was significantly reduced. Analysis of combined eye-head movements also revealed a greater number of poorly coordinated head movements in patients with schizophrenia, suggesting altered visuo-vestibular interaction. The authors attribute these differences to impairment of executive functions, typical of many neurological and psychiatric disorders, which compromises attentional processes and working memory necessary for the modulation of saccadic movements.

Notably, in schizophrenia, the same executive-function impairments that prevented enhancement of saccadic kinematics with increasing assigned salience also degraded the quality of visuo-vestibular interaction, further supporting the close relationship between visual attention, working memory, saccadic control, and visuo-vestibular integration. the study also showed that the effect of salience on voluntary saccadic kinematics is modulable, suggesting that this interaction may be amenable to targeted training.

While previous studies highlighted the relationship between visual attention, working memory, and visually guided voluntary saccades, Hawkins et al. (30) demonstrated similar effects for SHIMP saccades. Using vHIT, SHIMP responses were analyzed in patients with Parkinson’s disease and healthy controls. The authors proposed that the SHIMP saccade should be considered a subtype of visually induced voluntary saccade whose initiation also depends on vestibular and cervical inputs. A significant correlation was observed between cognitive decline, measured using the Mini-Mental State Examination (MMSE), and increased SHIMP saccadic latency in both patients and controls.

The observations of Kim E. Hawkins et al. (30) appear to find their anatomo-physiological substrate in the study by Liye Yi et al. (31), which shows that patients with mild cognitive impairment identified through MMSE present diffuse gray-matter atrophy and alterations in intrinsic functional brain activity. These changes were primarily localized in the superior and middle frontal gyri and in the prefrontal cortex, regions where neural circuits involved in visual attention, working memory, and oculomotor control converge. Together, these findings support the hypothesis of a close neuroanatomical and neurophysiological interaction between cognitive and oculomotor processes.

Furthermore, additional studies (32,33) have emphasized that alterations of associative cortical areas are associated with a significant reduction in recognition performance during functional video Head Impulse Testing (fHIT) in patients with Parkinson’s disease, vestibular migraine, and persistent postural-perceptual dizziness (PPPD), despite the presence of peripheral vestibular function within normal limits as assessed using the vHIT HIMP modality.

Additional experimental evidence (34) indicates that working memory and visual attention can significantly influence the dynamics of saccadic movements. In particular, saccadic latency is reduced when the visual target stimulus is associated with previously stored information, suggesting that cognitive processes of anticipation and recognition may modulate the efficiency of eye movements.

Overall, several experimental findings indicate that impairments of executive functions may compromise both the modulation of saccadic latency and visuo-vestibular coordination, highlighting the central role of cognitive processes in oculomotor control.

The review by Galit Yogev et al. (35) further demonstrated that reduced executive function significantly decreases gait speed and increases fall risk, particularly under dual-task conditions. Even in healthy young adults and children, performing a dual task during walking results either in reduced performance on the secondary task or reduced walking speed.

The present study, based on the simultaneous use of diagnostic tools commonly employed in the otoneurological field, aimed in the first two experiments to investigate the role of attentional processes and working memory in modulating the functional relationship between the horizontal canal slow phase and SHIMP saccades. To this end, a temporally divergent saccadic-interpretative dual-task was introduced to evaluate processing strategies within neural mechanisms shared by saccadic control and attentional processing when reactivation was required while processing of the previous task might still be ongoing.

In the third experiment, by contrast, the saccadic-interpretative dual-task was made temporally convergent in order to evaluate whether the interaction between saccadic function and recognition-related cognitive load could influence the latency and clustering of corrective saccades.

## 2. Materials and methods

### 2.1 Testing

#### 2.1.1 Mini-Mental State Examination (MMSE)

All participants underwent cognitive assessment using the Italian version of the MMSE. MMSE was conducted strictly face to face following the guidelines and protocols by investigators and was completed during 5–10 min. The MMSE is a 30-point questionnaire used extensively in clinical and research settings to measure cognitive impairment, including simple tasks in a number of areas: the test of time and place, the repeating lists of words, arithmetic such as serial subtractions of seven, language use and comprehension, and basic motor skills.

#### 2.1.2 Dizziness Handicap Inventory (DHI)

The dizziness handicap inventory (DHI) was created by Jacobson & Newman to evaluate the quality of life and its impact on daily functioning for patients suffering from vertigo and dizziness. It is composed of 25 items, with three main subgroups including functional, emotional, and physical aspects of dizziness and unsteadiness. DHI is widely used as a self-reported measurement of dizziness. The DHI was self-completed by each patient before vestibular testing.

#### 2.1.3 Video Head Impulse test (vHIT)

All subjects were assessed with the vHIT to measure the VOR gain for each semicircular canal. VOR gain for horizontal (HSC), superior (SSC), and posterior semicircular canals (PSC) in response to high-frequency head stimuli was tested using an ICS video-oculographic system (GN Otometrics, Taastrup, Denmark).

Passive, unpredictable 5°–20°, 50°–250°/s, and 750°–5000°/s^2^ head impulses were delivered manually on the plane of the horizontal and vertical canals (RALP and LARP planes) while the patient was asked to keep looking at an earth-fixed target. At least 15 stimuli were delivered for stimulating each canal and averaged to obtain the corresponding mean VOR gain. VOR gain values <0.8 for HSC and <0.7 for SSC and PSC with corrective saccades (overt and/or covert) were considered pathological.

#### 2.1.4 Cervical and ocular vestibular-evoked myogenic potentials

Cervical and ocular vestibular-evoked myogenic potentials (cVEMPs and oVEMPs, respectively) for air-conducted sounds were recorded using 2-channel evoked potential acquisition systems (either Neuro-Audio, Neurosoft, Russia or Viking, Nicolet EDX, CareFusion, Germany depending on different centers) with surface electrodes placed according to standardized criteria. Potentials were recorded delivering tone bursts (frequency: 500 Hz, duration: 8 ms, stimulation rate: 5 Hz) via headphones either before or following CRP. The recording system used an EMG-based biofeedback monitoring method to minimize variations in muscle contractions and VEMPs amplitudes. A re-test was performed for each stimulus to assess reproducibility. The first biphasic responses on the ipsilateral sternocleidomastoid muscle (p13-n23) for cVEMPs (ipsilateral response) and under the patient’s contralateral eye (n10-p15) for oVEMPs (crossed response) were analyzed by calculating the peak-to-peak amplitude.

#### 2.1.5 Functional Head Impulse Test (fHIT)

The fHIT was performed using the fHIT system (Beon Solutions srl, Zero Branco (TV), Italy). Patients were seated in a static chair in front of a computer screen at a distance of 1.5 meter with a keyboard in their hand. During a passive head impulse, when head acceleration reached its peak value, an optotype (Landolt C ring) was displayed on the screen for 80 ms. The size of the optotype was adjusted for every subject separately, and remained constant during testing. Before the start of the fHIT, the static visual acuity threshold was acquired by the fHIT system in 20 trials. Optotype size started from 1.0 LogMAR (log of the Minimum Angle of Resolution) and decreased depending on the subjects’ rates of errors. The used optotype size was equal to this threshold, increased by 0.6 LogMAR. During fHIT, patients had to choose the right optotype out of eight different options by pressing the corresponding button on the keyboard. No direct feedback was given. Head impulses comprised fast (peak velocity >150°/s), outwards, passive, horizontal rotational head movements with a low amplitude (±20°), unpredictable in timing and direction. At least 10 impulses were given to both sides. The absolute outcome was the percentage of correct answers (%CA) for each side, as calculated by the fHIT system. A %CA of <80 was considered abnormal. This cut-off was a conservative approximation of the criterion adopted by the fHIT system, which considers the level where the standardized normal deviate of the patient falls outside the 99% of the two-tailed Z distribution of a population of age-matched controls.

#### 2.1.6 Patient setup considerations for simultaneous vHIT–fHIT testing

Simultaneous acquisition of vHIT and fHIT can be performed without introducing recording artifacts only in subjects whose cranial morphology allows the concurrent use of the vHIT goggles and the fHIT headband without physical contact or overlap between the two devices. This anatomical requirement inevitably limited the number of subjects eligible for inclusion in the three experiments. The present retrospective study included participants who had been recruited between May 2020 and April 2021.

Prior to the simultaneous recordings, all participants underwent standard SHIMP testing and standard fHIT testing, which served both as baseline measurements and to facilitate familiarization with the two procedures. Participants were then positioned in front of the fHIT display, which was necessarily large enough to ensure that the vHIT laser target (SHIMP protocol) remained within the monitor boundaries during head rotations. After a new assessment of visual acuity, a central white fixation point appeared on the screen and was rapidly replaced by the optotype at the peak head-rotation velocity. Participants were instructed to fixate the central point while keeping the head still during placement of the vHIT goggles. Calibration of the vHIT system was then performed by aligning the white fixation point on the monitor between the two red laser targets of the system, following the same calibration procedure used in conventional vHIT testing.

In the first two experiments, which involved the SHIMP protocol, participants were explicitly instructed to prioritize execution of the SHIMP saccade. Specifically, they were asked to report identification of the optotype using the keyboard only after reacquiring the red laser target as quickly as possible immediately following the head rotation, in order to prevent the interpretative processing phase from coinciding with, and potentially interfering with, the correct execution of the anticompensatory saccade.

All examinations related to the three experiments described below were performed on the same day, within a few minutes, following the initial baseline evaluation.

### 2.2 Procedure

#### 2.2.1 Experiment 1

A sample of ten healthy participants (5 males and 5 females; age range: 34–63 years), without vestibular disorders or cognitive impairment (see Table A), was recruited. Each participant underwent a complete otoneurological assessment including Mini-Mental State Examination (MMSE), comprehensive bedside examination, vHIT testing in both HIMP and SHIMP modes, ocular and cervical vestibular evoked myogenic potentials (oVEMPs and cVEMPs), and Functional Head Impulse Test (fHIT). The set of these investigations, performed separately, was defined as the “separate protocol”.

**Table A:** Results from the first experiment comparing separate and combined testing protocols (vHIT–SHIMP paradigm and fHIT) in 10 healthy participants.

| PARTICIPANT | SEX | MMSE SCORE | SEPARATE PROTOCOL-SHIMP |  |  |  |  |  |  |  | SEPARATE PROTOCOL-FHIT |  | COMBINED PROTOCOL-SHIMP |  |  |  |  |  |  |  | COMBINED PROTOCOL-FHIT |  |
| --- | --- | --- | --- | --- | --- | --- | --- | --- | --- | --- | --- | --- | --- | --- | --- | --- | --- | --- | --- | --- | --- | --- |
|  |  |  | GAIN LEFT | GAIN RIGHT | MEAN LATENCY LEFT | MEAN LATENCY RIGHT | PR LEFT | PR RIGHT | AMPLITUDE LEFT | AMPLITUDE RIGHT | % FUNCTIONAL LEFT | % FUNCTIONAL RIGHT | GAIN LEFT | GAIN RIGHT | MEAN LATENCY LEFT | MEAN LATENCY RIGHT | PR LEFT | PR RIGHT | AMPLITUDE LEFT | AMPLITUDE RIGHT | % FUNCTIONAL LEFT | % FUNCTIONAL RIGHT |
| PARTICIPANT 1 | F | 29 | 0.84 | 0.95 | 168 | 167 | 26 | 32 | 405 | 349 | 100 | 100 | 0.91 | 0.94 | 399 | 376 | 38 | 30 | 374 | 286 | 100 | 87 |
| PARTICIPANT 2 | F | 29 | 0.89 | 1.02 | 182 | 249 | 37 | 74 | 308 | 239 | 100 | 100 | 0.93 | 1.02 | 364 | 346 | 51 | 42 | 328 | 297 | 89 | 95 |
| PARTICIPANT 3 | M | 28 | 0.87 | 1.04 | 185 | 196 | 26 | 43 | 336 | 380 | 100 | 100 | 1 | 1.01 | 439 | 432 | 24 | 30 | 288 | 359 | 88 | 90 |
| PARTICIPANT 4 | M | 26 | 0.82 | 0.96 | 272 | 220 | 42 | 46 | 373 | 379 | 100 | 100 | 0.82 | 0.91 | 350 | 339 | 53 | 30 | 329 | 322 | 77 | 80 |
| PARTICIPANT 5 | F | 27 | 0.82 | 0.92 | 174 | 168 | 25 | 31 | 309 | 305 | 100 | 100 | 0.77 | 0.92 | 189 | 224 | 43 | 24 | 293 | 319 | 68 | 68 |
| PARTICIPANT 6 | M | 27 | 0.85 | 0.91 | 175 | 170 | 22 | 30 | 290 | 300 | 100 | 100 | 0.9 | 0.84 | 350 | 320 | 45 | 50 | 288 | 305 | 94 | 90 |
| PARTICIPANT 7 | F | 27 | 0.87 | 0.9 | 183 | 200 | 33 | 40 | 370 | 379 | 98 | 98 | 0.85 | 0.85 | 330 | 390 | 60 | 62 | 380 | 380 | 95 | 94 |
| PARTICIPANT 8 | M | 29 | 1 | 0.9 | 170 | 180 | 42 | 43 | 290 | 300 | 100 | 100 | 0.98 | 0.95 | 350 | 340 | 55 | 60 | 300 | 278 | 100 | 100 |
| PARTICIPANT 9 | F | 28 | 0.85 | 0.82 | 168 | 180 | 25 | 30 | 320 | 332 | 100 | 100 | 0.87 | 0.92 | 400 | 399 | 50 | 45 | 360 | 380 | 100 | 100 |
| PARTICIPANT 10 | M | 29 | 1 | 0.99 | 190 | 200 | 33 | 48 | 329 | 378 | 100 | 100 | 0.9 | 0.92 | 343 | 355 | 50 | 63 | 360 | 362 | 90 | 90 |

The analysis focused on mean latency and degree of clustering of SHIMP saccades (PR index), as well as on optotype recognition performance during the fHIT. Subsequently, the same participants underwent simultaneous vHIT testing in SHIMP mode and fHIT testing on the lateral semicircular canals. This procedure was defined as the “combined protocol”. During this condition, participants were instructed to prioritize execution of the SHIMP saccade and only afterward report the optotype perceived during the head rotation.

This experimental paradigm was designed to induce a temporary competition between cognitive processes, specifically between visual attention and working memory required for optotype recognition, and the voluntary saccadic movement required by the SHIMP protocol. The aim was to determine whether healthy participants were able to perform both tasks simultaneously without alterations in visuo-vestibular interaction (evaluated through the temporal relationship between the VOR slow phase, mean latency, and clustering of SHIMP saccades) and without reductions in optotype recognition performance (Landolt C), or whether a functional trade-off emerged between oculomotor performance and cognitive task execution, similar to that observed in dual-task paradigms during gait (35).

#### 2.2.2 Experiment 2

A group of ten cognitively intact patients with previous unilateral or bilateral vestibular deficits in the chronic phase was recruited (see Table B). These patients were characterized by the presence of SHIMP saccades on the affected lateral semicircular canal and by residual symptoms ranging from mild to disabling according to the Dizziness Handicap Inventory (DHI).

**Table B:** Clinical characteristics of patients with vestibular disorders (see diagnosis column) and corresponding DHI scores. Data from both separate and combined SHIMP–fHIT protocols are reported, including lateral semicircular canal gains, bilateral mean saccadic latency, PR score, mean saccadic amplitude, and percentage of correct responses on the fHIT. ( NA = Not applicable)

|  |  |  |  | SEPARATE PROTOCOL-SHIMP |  |  |  |  |  |  |  | SEPARATE PROTOCOL-FHIT |  | COMBINED PROTOCOL-SHIMP |  |  |  |  |  |  |  | COMBINED PROTOCOL-FHIT |  |
| --- | --- | --- | --- | --- | --- | --- | --- | --- | --- | --- | --- | --- | --- | --- | --- | --- | --- | --- | --- | --- | --- | --- | --- |
| PATIENT | SEX | DIAGNOSIS | DHI SCORE | GAIN LEFT | GAIN RIGHT | MEAN LATENCY LEFT | MEAN LATENCY RIGHT | PR LEFT | PR RIGHT | AMPLITUDE LEFT | AMPLITUDE RIGHT | % FUNCTIONAL LEFT | % FUNCTIONAL RIGHT | GAIN LEFT | GAIN RIGHT | MEAN LATENCY LEFT | MEAN LATENCY RIGHT | PR LEFT | PR RIGHT | AMPLITUDE LEFT | AMPLITUDE RIGHT | % FUNCTIONAL LEFT | % FUNCTIONAL RIGHT |
| PATIENT 1 | M | RIGHT AVD | 40 | 0.84 | 0.67 | 197 | 201 | 85 | 54 | 259 | 199 | 100 | 69 | 0.92 | 0.6 | 352 | ABSENT | 52 | NA | 216 | ABSENT | 100 | 64 |
| PATIENT 2 | F | RIGHT AVD | 70 | 0.8 | 0.38 | 259 | 283 | 34 | 24 | 314 | 197 | 70 | 25 | 0.72 | 0.33 | 405 | ABSENT | 31 | NA | 260 | ABSENT | 85 | 6.3 |
| PATIENT 3 | M | RIGHT AVD | 12 | 0.91 | 0.69 | 195 | 300 | 39 | 46 | 431 | 304 | 100 | 83 | 0.91 | 0.6 | 357 | 400 | 51 | 32 | 390 | 325 | 93.9 | 50 |
| PATIENT 4 | F | PRESBYVE ST | 20 | 0.82 | 0.91 | 181 | 197 | 19 | 18 | 417 | 410 | 100 | 100 | 0.85 | 0.87 | 269 | 284 | 40 | 58 | 378 | 333 | 91 | 66 |
| PATIENT 5 | F | RIGHT AVD | 10 | 0.82 | 0.6 | 193 | 203 | 16 | 73 | 292 | 236 | 100 | 90.9 | 0.81 | 0.65 | 218 | 176 | 32 | 43 | 337 | 326 | 66 | 26 |
| PATIENT 6 | M | LEFT AVD | 18 | 0.58 | 0.9 | 180 | 210 | 35 | 30 | 190 | 260 | 55 | 100 | 0.5 | 0.85 | 450 | 350 | 40 | 45 | 100 | 280 | 25 | 90 |
| PATIENT 7 | F | RIGHT AVD | 50 | 0.85 | 0.45 | 168 | 250 | 41 | 35 | 261 | 200 | 95 | 28 | 0.83 | 0.47 | 350 | ABSENT | 58 | NA | 270 | ABSENT | 100 | 10 |
| PATIENT 8 | M | RIGHT AVD | 18 | 0.9 | 0.6 | 210 | 280 | 35 | 48 | 230 | 210 | 100 | 75 | 0.85 | 0.62 | 380 | 530 | 45 | 72 | 245 | 200 | 94 | 38 |
| PATIENT 9 | M | RIGHT AVD | 54 | 0.95 | 0.53 | 201 | 197 | 48 | 53 | 180 | 130 | 90 | 53 | 0.9 | 0.6 | 380 | ABSENT | 42 | NA | 190 | ABSENT | 95 | 45 |
| PATIENT 10 | F | BVD | 70 | 0.48 | 0.5 | 240 | 230 | 53 | 48 | 130 | 140 | 48 | 40 | 0.51 | 0.48 | ABSENT | ABSENT | NA | NA | ABSENT | ABSENT | 15 | 10 |

In these participants, the combined protocol (vHIT–SHIMP performed simultaneously with fHIT) was applied in order to evaluate whether the interaction between the two instruments could reveal deterioration of SHIMP saccadic parameters on both the affected and contralateral sides when patients were required to perform a visual recognition task nearly simultaneously with the execution of anticompensatory saccades.

This paradigm was designed to reproduce, under experimental conditions, a visuo-vestibular and cognitive dual-task scenario similar to complex real-life situations in which patients must maintain adequate visuo-vestibular stability while simultaneously processing multiple visual inputs necessary for correct interpretation of the surrounding environment. Such conditions frequently occur in natural ecological contexts and are particularly relevant during gait; however, this situation could not be experimentally reproduced in the present study.

It was also evaluated whether any worsening of mean latency and clustering of SHIMP saccades observed during the combined protocol was more pronounced than in healthy participants and whether such alterations showed a correlation with DHI scores.

#### 2.2.3 Experiment 3

An additional group of ten cognitively intact patients with previous unilateral or bilateral vestibular deficits was recruited (see Table Ca and Cb). These participants underwent the same vestibulometric assessment protocol adopted in the previous experiments.

In this third protocol, however, the fHIT was performed simultaneously with vHIT testing in HIMP mode rather than with the SHIMP protocol. Participants were instructed to maintain gaze at the center of the monitor during head rotations while simultaneously attempting to recognize the optotype presented during the stimulus, in order to introduce a cognitive task associated with the execution of corrective saccades.

The aim was to evaluate a possible improvement in mean latency and clustering of corrective saccades within a temporally convergent dual-task condition. Head impulses toward the affected side were initially delivered at low velocity, allowing participants to recognize the optotype despite reduced VOR gain; stimulus velocity was then unpredictably increased to appropriate values while monitoring the saccadic response using vHIT.

This stimulation strategy was based on the assumption that “persistent activity”, a term often considered synonymous with working memory, does not exclusively reflect maintenance of previously encoded information but is also involved in anticipation of imminent actions by representing motor goals during the preparatory interval preceding an expected stimulus (36). This approach justified the initial administration of low-velocity head impulses toward the affected side, allowing activation of symbolic processing mechanisms despite reduced VOR gain. In this way, the cognitive effort directed toward recognition was effectively integrated into the experimental protocol.

An alternative methodological approach could have involved asking participants, during presentation of the central fixation point, to mentally represent a previously indicated Landolt C optotype and subsequently determine during head rotation whether the optotype presented matched or differed from the imagined stimulus. However, this option was excluded because it relied excessively on abstraction abilities that could vary considerably between participants.

## 3. Results

### 3.1 Experiment 1

#### 3.1.1 Sample

Ten healthy participants (5 males, 5 females), aged 34–63 years, without vestibular disorders or cognitive impairment (MMSE > 25), were included.

Participants underwent evaluation using both the separate protocol and the combined protocol.

#### 3.1.2 Parameters measured

The following parameters were recorded:

- VOR gain of the lateral semicircular canals using vHIT in SHIMP mode (separate protocol);
- Presence or absence of SHIMP saccades in the lateral canals (separate protocol);
- Mean latency of SHIMP saccades for both lateral canals (separate protocol);
- Clustering of SHIMP saccades expressed as the PR index for both lateral canals (separate protocol);
- Mean amplitude of SHIMP saccades for both lateral canals (separate protocol);
- Percentage of Landolt C optotype recognition at the Functional Head Impulse Test (fHIT) (separate protocol).

All six parameters listed above were subsequently recorded again in the same participants using the combined protocol (vHIT–SHIMP combined with fHIT). The results of Experiment 1 are summarized in Table A.

As shown in Table A, the only parameters that demonstrated significant variation when comparing the separate protocol with the combined protocol were the mean latency of SHIMP saccades and the percentage of optotype recognition at the fHIT.

In particular, mean latency nearly doubled, with an average increase of approximately 85% (estimated range: +62% to +108%), showing high statistical significance (p = 0.00001798075888).

In contrast, the degree of saccadic clustering, expressed by the PR index, showed a mean increase of 29.06%, with borderline statistical significance (p = 0.04876459867) and an estimated variation range between +6.0% and +52.1%, indicating a statistically significant result characterized by moderate uncertainty.

No statistically significant changes were observed in saccadic amplitude (p-value=0,6935596028) and VOR gains (p-value left side=0,5743560191, p-value right side= 0,4689535051).

The percentage of optotype recognition at fHIT showed statistically significant differences (p = 0.01058223221), indicating that the negative effect observed under the simultaneous protocol was unlikely to be attributable to chance.

In 8 out of 10 participants, optotype recognition performance remained substantially unchanged between the two protocols, whereas the mean latency of SHIMP saccades increased during the combined protocol.

In the remaining two participants: one showed a smaller increase in mean latency (approximately 40%), accompanied by reduced optotype recognition performance (from 100% bilaterally under the separate protocol to 80% right and 77% left under the combined protocol), the other showed stable saccadic latency, but a marked decrease in optotype recognition performance (from 100% bilaterally to 68% bilaterally).

Overall, these findings suggest that the near-simultaneous presentation of two cognitive demands induces an implicit allocation of visuo-vestibular-attentional resources in healthy participants.

In 80% of cases, this allocation favored correct processing of the visual stimulus at the expense of a partial deterioration in the interaction between the vestibulo-ocular reflex and SHIMP saccades, as reflected by the increase in mean latency.

This deterioration was bilateral and symmetrical and affected only mean latency, without substantially modifying saccadic clustering (PR index).

In other words, under dual-task conditions, healthy participants appear to require longer processing time before initiation of SHIMP saccades, while maintaining a relatively preserved temporal organization of anticompensatory saccades, as indicated by the relative stability of the PR index.

This strategy allows preservation of Landolt C recognition performance, despite a modest slowing of the oculomotor response which, given the stability of the PR index, does not result in disruption of visuo-vestibular interaction.

Representative examples are reported below.

**Figure 1:**
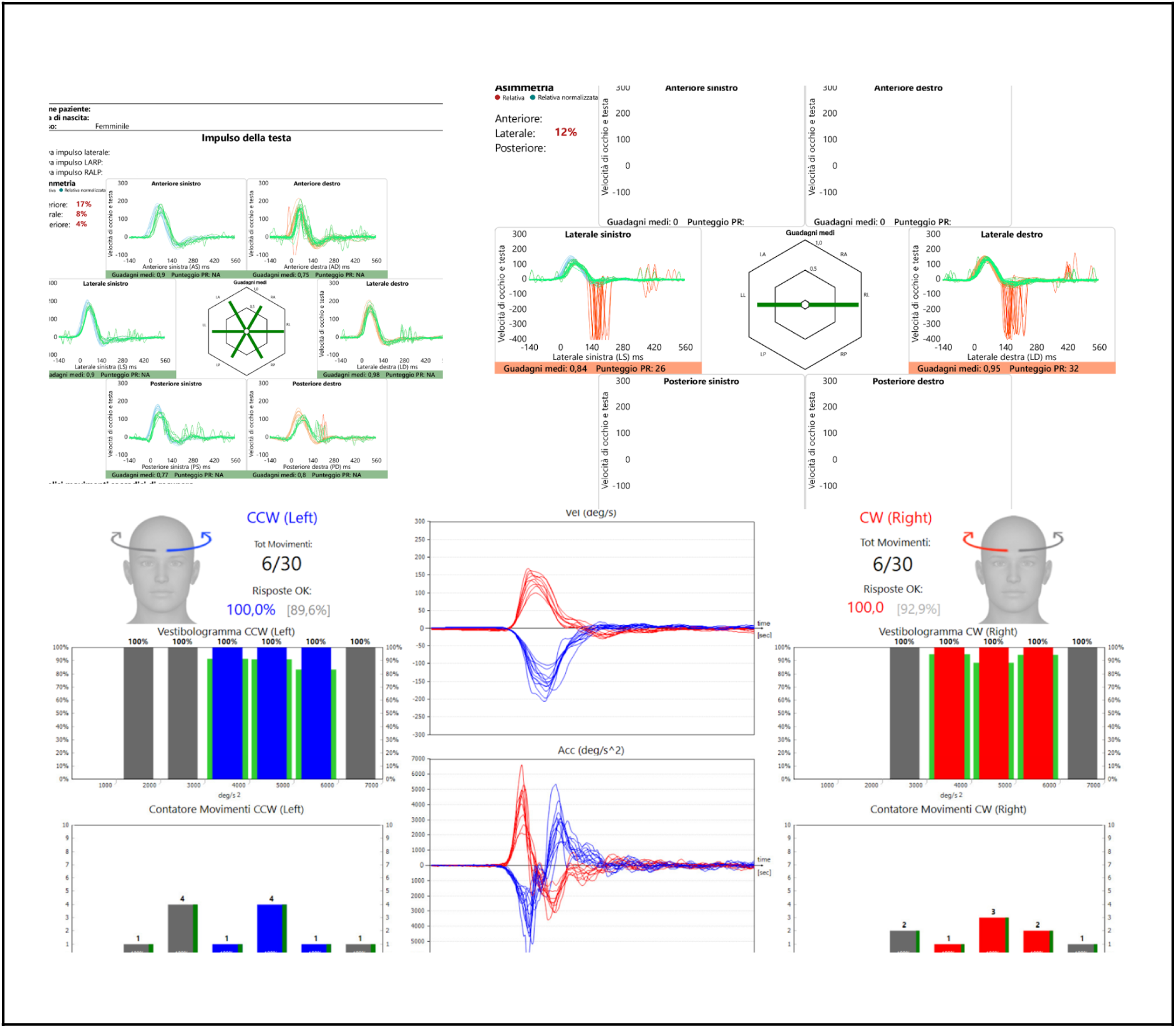
Representative normal results obtained with the separate testing protocol. vHIT showed normal gains in all six semicircular canals, with physiological SHIMP responses and normal fHIT performance.

**Figure 2:**
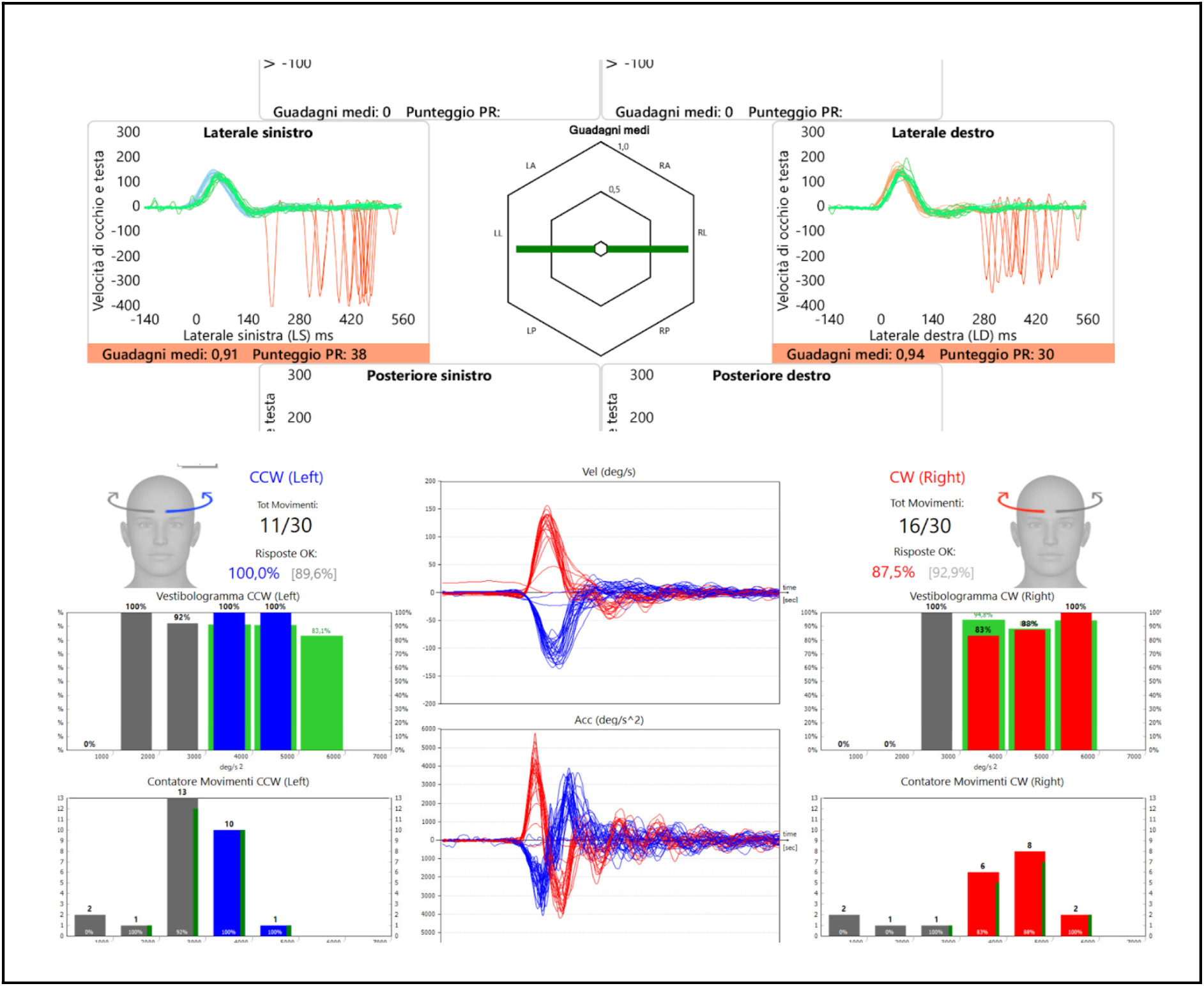
Results from the same participant during the combined testing protocol. SHIMP responses demonstrated increased saccadic latency bilaterally, with a reduction in the percentage of correct responses on the right side at the fHIT.

A marked increase in mean SHIMP saccadic latency was observed when moving from the separate protocol to the combined protocol, more evident on the right side. Mean latency increased from 168 ms (left) and 167 ms (right) under the separate protocol to 399 ms (left) and 376 ms (right) under the combined protocol.

In contrast, the degree of saccadic clustering, expressed by the PR index, remained substantially stable between protocols: 26 (left) and 32 (right) under the separate protocol, 38 (left) and 30 (right) under the combined protocol.

These findings indicate a generalized delay in SHIMP saccade initiation under dual-task conditions, with only limited changes in the temporal distribution of saccadic latencies.

Optotype recognition performance was also partially influenced by the dual-task condition. Recognition remained 100% on the left side, whereas a reduction to 87% on the right side was observed, suggesting mild deterioration in visual performance associated with increased cognitive and oculomotor demands.

**Figure 3:**
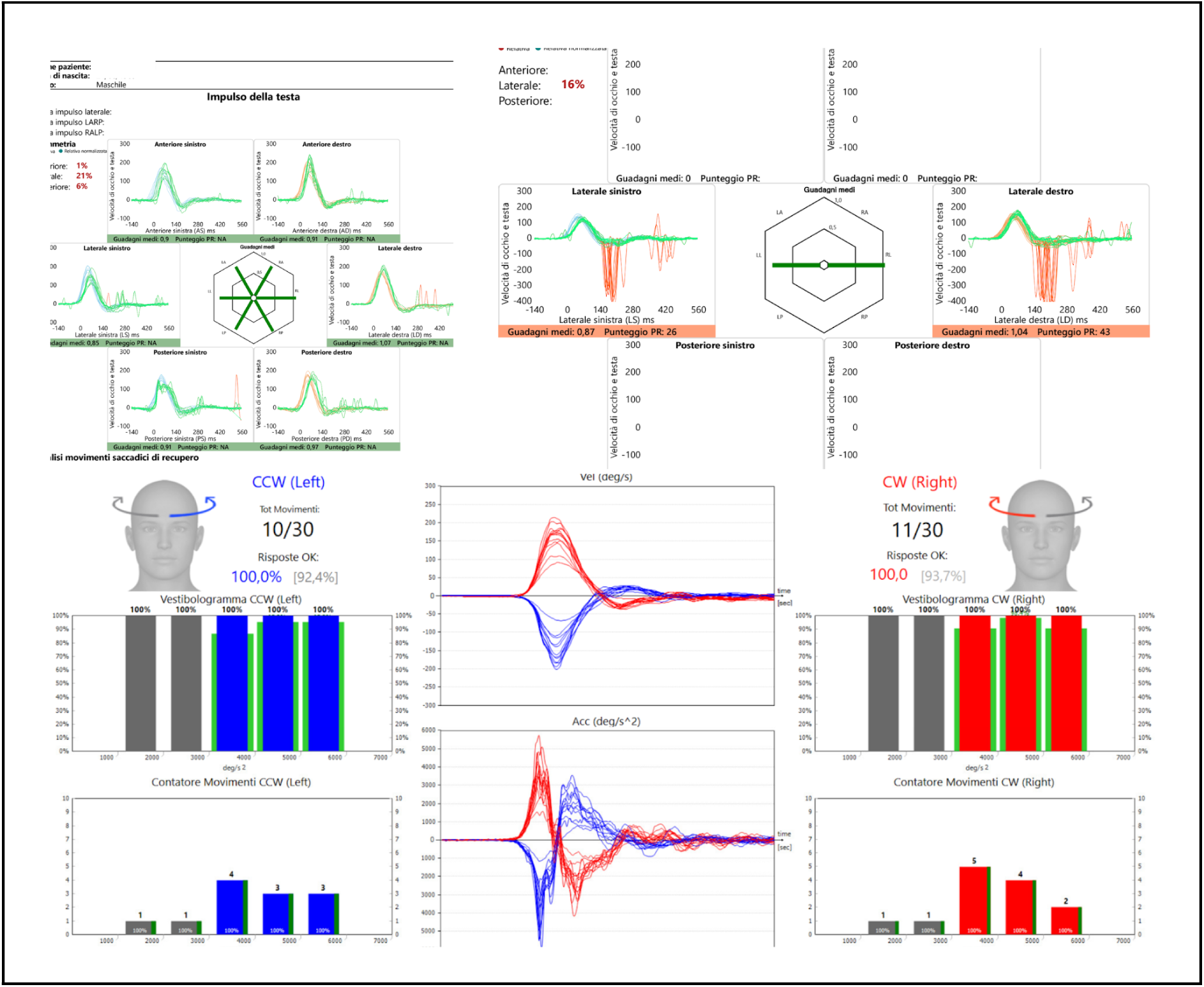
Normal results at the separate testing protocol. vHIT gains were within normal limits in all six semicircular canals, with physiological SHIMP responses and normal performance on the fHIT.

**Figure 4:**
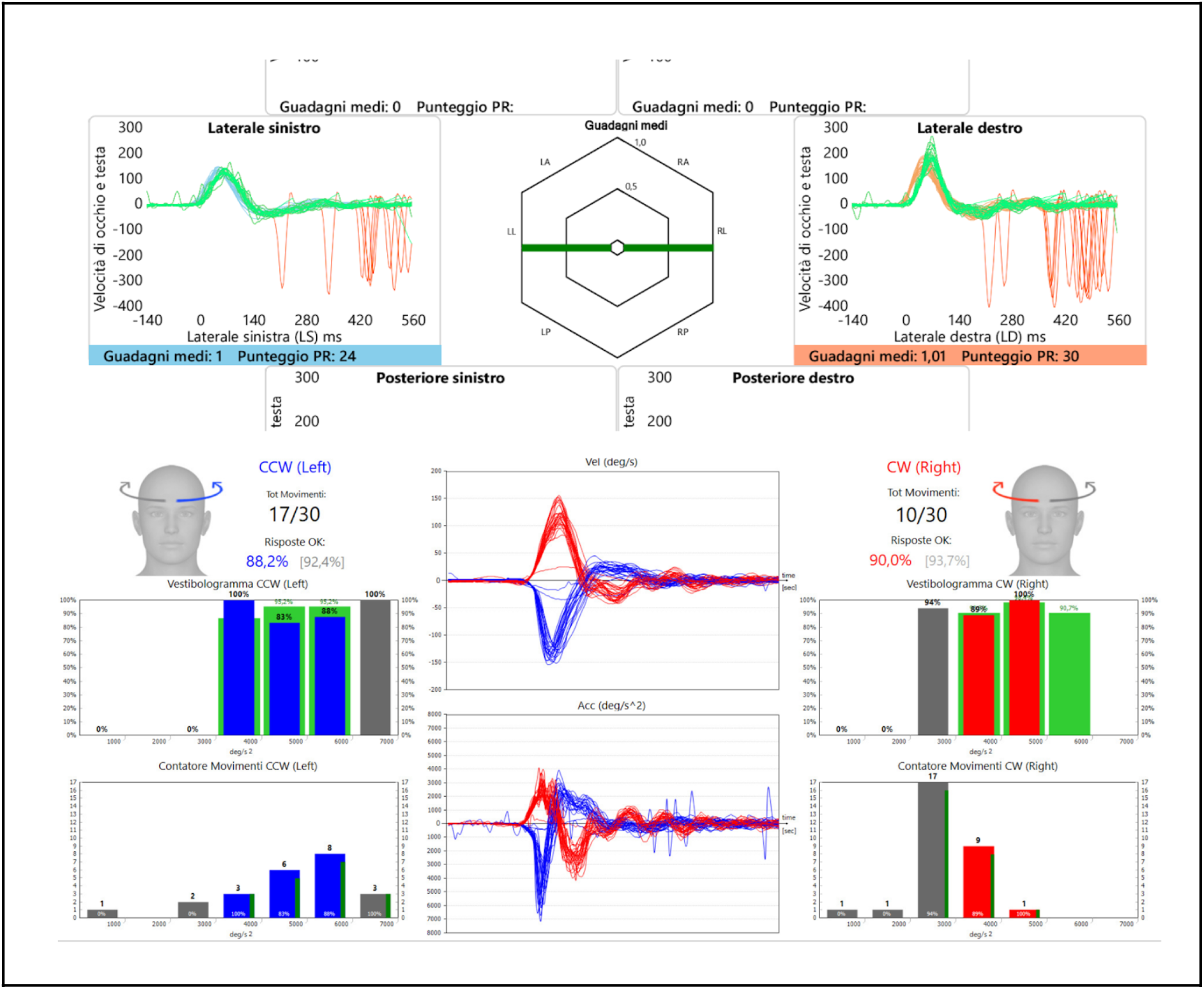
Results from the same participant during the combined testing protocol. SHIMP responses showed increased saccadic latency bilaterally and reduced percentage of correct responses on the fHIT.

A marked increase in mean SHIMP saccadic latency was again observed under the combined protocol. Mean latency increased from 185 ms (left) and 196 ms (right) to 439 ms (left) and 432 ms (right). In contrast, saccadic clustering showed only minimal changes from 26 (left) and 43 (right) under the separate protocol to 24 (left) and 30 (right) under the combined protocol.

Overall, these data confirm a significant delay in SHIMP saccade initiation without relevant alterations in temporal organization of saccadic responses.

A partial reduction in optotype recognition performance was also observed, decreasing to 88% on the left side and 90% on the right side.

### 3.2 Experiment 2

#### 3.2.1 Sample

Ten patients (5 males, 5 females), aged 28–73 years, with previous vestibular insults of different etiologies were recruited.

#### 3.2.2 Clinical distribution

- 6 patients with right superior vestibular nerve branch acute vestibular deficit (AVD)
- 1 patient with complete right-sided AVD
- 1 patient with left superior vestibular nerve branch AVD
- 1 patient with presbyvestibulopathy (DHI = 20)
- 1 patient with marked bilateral vestibular hyporeflexia (DHI = 70)

All patients were in the chronic phase, approximately one year after the acute event. Dizziness Handicap Inventory (DHI) scores ranged from 12 to 70 (See Table B).

#### 3.2.3 Parameters measured (separate protocol)

The following parameters were recorded:

- VOR gain of the lateral semicircular canals using vHIT in SHIMP mode
- Presence or absence of SHIMP saccades in the lateral canals
- Mean latency of SHIMP saccades for both lateral canals
- Clustering of SHIMP saccades expressed as the PR index for both lateral canals
- Mean amplitude of SHIMP saccades for both lateral canals
- Percentage of Landolt C optotype recognition at the Functional Head Impulse Test (fHIT)
- Subjective disability assessed using the Dizziness Handicap Inventory (DHI)

The first six parameters listed above were subsequently recorded again in the same participants using the combined protocol (vHIT–SHIMP combined with fHIT).

In the second patient group, similarly to what was observed in the healthy control group of Experiment 1, the parameters most affected by the transition from the separate protocol to the simultaneous protocol were mean SHIMP saccadic mean latency and Landolt C recognition performance at fHIT. No statistically significant changes were observed in VOR gains (p-value left side=0,3846430173, p-value right side= 0,3679149188).

However, compared with healthy participants, a distinctive finding emerged: in 5 out of 10 patients, SHIMP saccades that were present during the separate protocol completely disappeared during the combined protocol. Notably, in patients with higher DHI scores (>40), anticompensatory saccades were absent throughout the sample under simultaneous testing conditions.

Among the eight patients with sequelae of acute vestibular deficit (AVD), four individuals who showed clearly detectable SHIMP saccades on the affected side during the separate protocol no longer exhibited them during the combined protocol. The same phenomenon was also observed in patient with bilateral vestibular areflexia. All these patients presented DHI scores between 40 and 70, consistent with moderate-to-severe disability.

In contrast, three patients with lower DHI scores (10–18) showed a different pattern: mean SHIMP saccadic latency increased by approximately 59% during the combined protocol compared with the separate protocol, reaching statistical significance (p = 0.0384387578), suggesting a likely genuine effect, although with some uncertainty in the precision of the estimate.

In this subgroup changes in the PR index were not statistically significant (p-value=0,5027977557) and no significant changes were observed in saccadic amplitude (p-value= 0,5889190711).

However, this relative preservation of visuo-vestibular dynamics occurred at the expense of optotype recognition performance, with marked reductions in fHIT identification accuracy.

Overall, the reduction in recognition performance during the simultaneous protocol was highly statistically significant (p = 0.0001578524536), with a mean decrease of approximately 47%, representing a robust negative effect that reached approximately 51% when considering only patients with DHI scores >40.

Specifically, recognition performance decreased:

- from 83% to 50% in one patient
- from 55% to 25% in another
- from 90% to 26% in a third patient

In these same subjects, lateral canal gain on the affected side measured with simple SHIMP testing was closer to normal values than in patients with higher DHI scores.

The patient with presbyvestibulopathy showed a marked increase in SHIMP saccadic latency during the combined protocol (from 181 ms left and 197 ms right under the separate protocol to 378 ms left and 333 ms right under the combined protocol), associated with reduced recognition performance (from 100% bilaterally under the separate protocol to 91% left and 66% right under the combined protocol).

Similarly, the patient with bilateral vestibular areflexia, in whom SHIMP saccades were completely absent during the combined protocol, also showed a dramatic deterioration in fHIT performance, decreasing from 40% bilaterally to 10% right and 15% left.

Representative examples of instrumental data are reported below.

**Figure 5:**
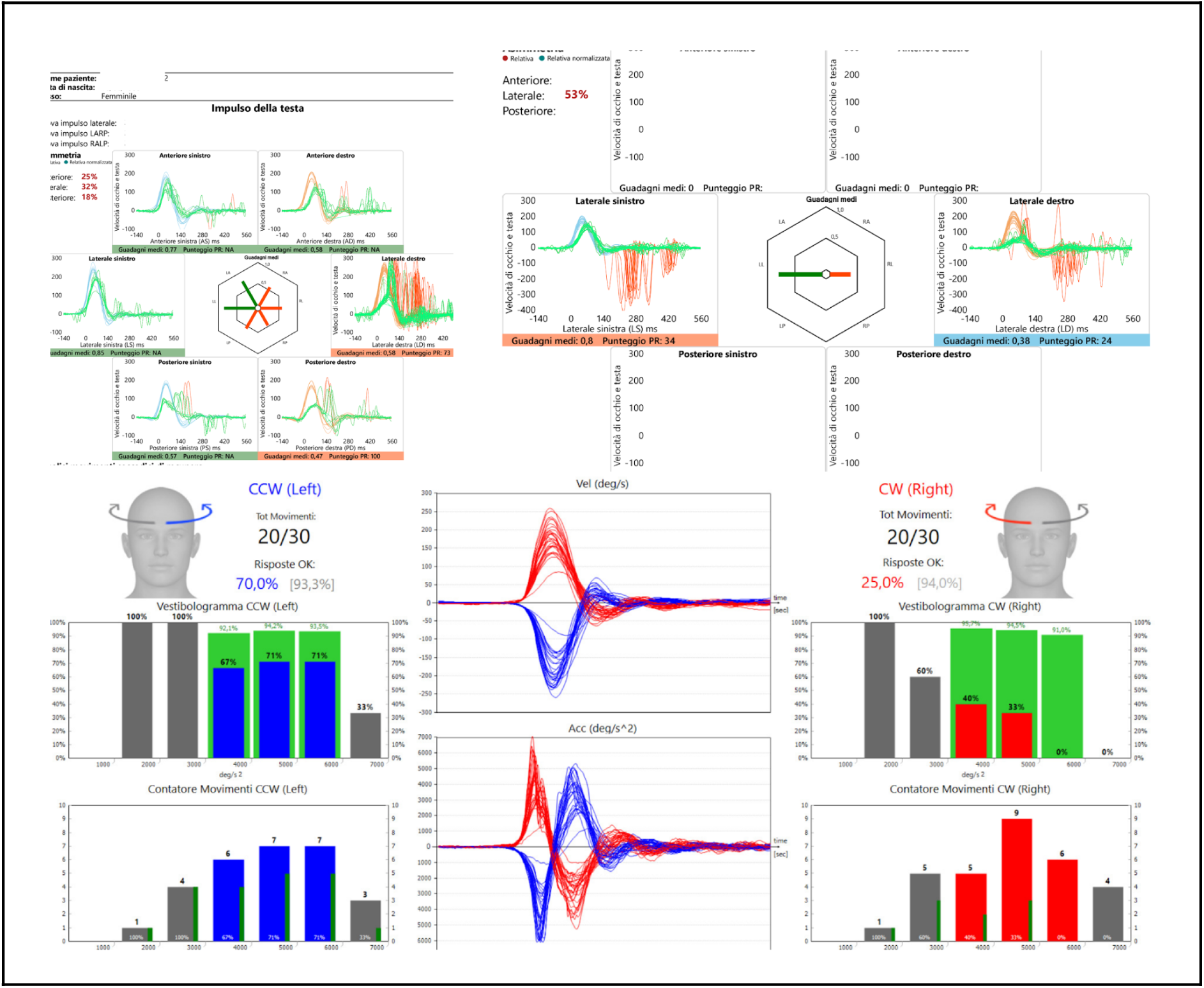
Representative traces obtained with the separate testing protocol. vHIT in the HIMP paradigm demonstrated a complete right-sided vestibular deficit. SHIMP responses showed reduced gain with anticompensatory saccades. fHIT performance was reduced, particularly on the right side (25%).

**Figure 6:**
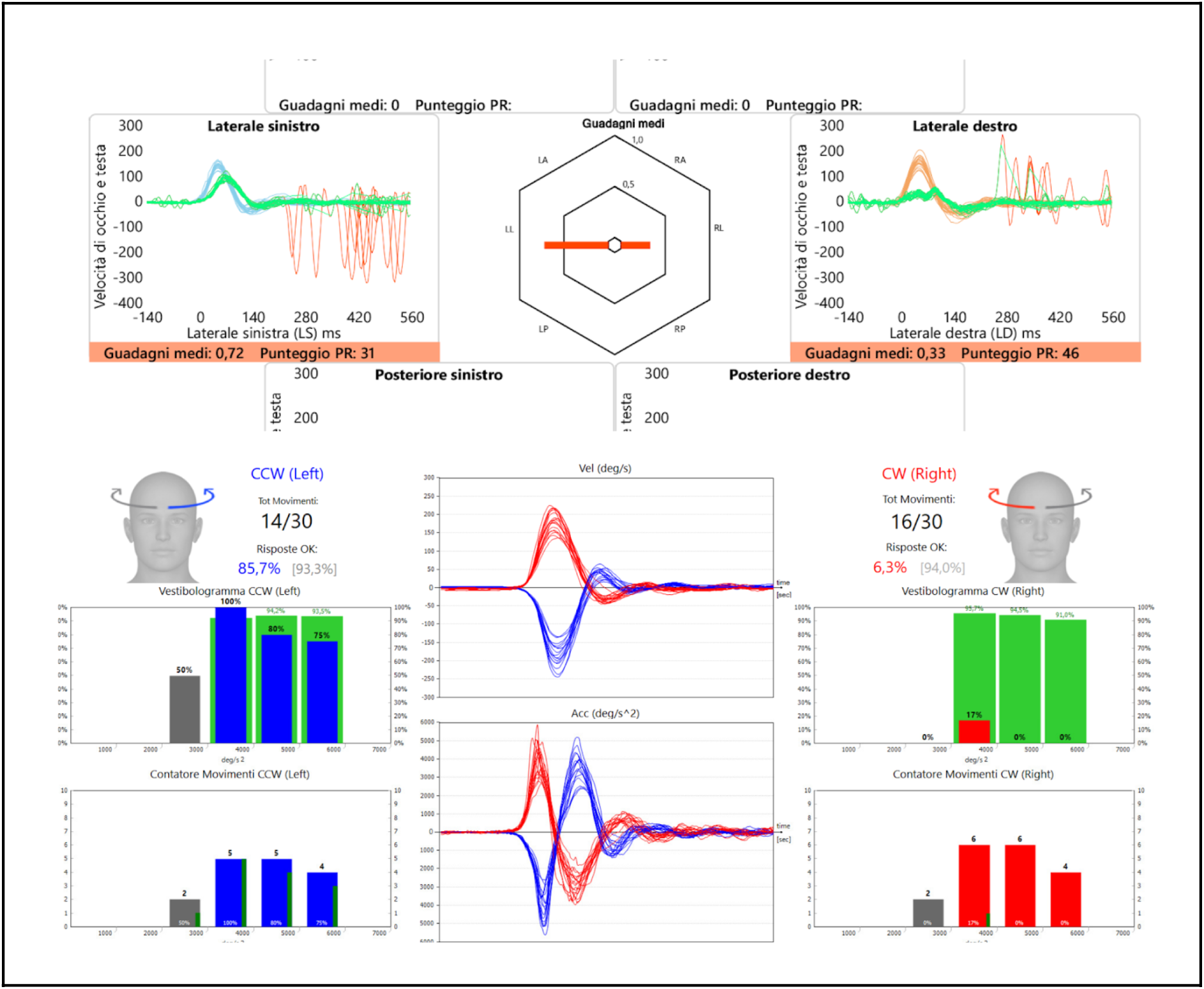
Results obtained during the combined testing protocol. The upper panel shows SHIMP responses, demonstrating a marked increase in anticompensatory saccadic latency on the left side and their complete absence on the right side. The lower panel shows fHIT results, with a substantial reduction in the percentage of correct responses during rightward head rotations, while leftward rotations remained unaffected.

Instrumental recordings obtained under the separate protocol showed preserved SHIMP saccades on the affected side.

Under the combined protocol, a persistent and complete disruption of visuo-vestibular interaction on the right side was observed during concurrent presentation of the cognitive task. Under these conditions, SHIMP saccades disappeared on the affected side, indicating marked interference of cognitive load with previously acquired vestibulo-oculomotor compensatory mechanisms.

At the same time, a significant reduction in recognition-task performance toward the affected side was observed, with correct-response rates decreasing from 25% under the separate protocol to approximately 6% under the combined protocol, suggesting a pronounced cognitive-vestibular interference effect under dual-task conditions.

**Figure 7:**
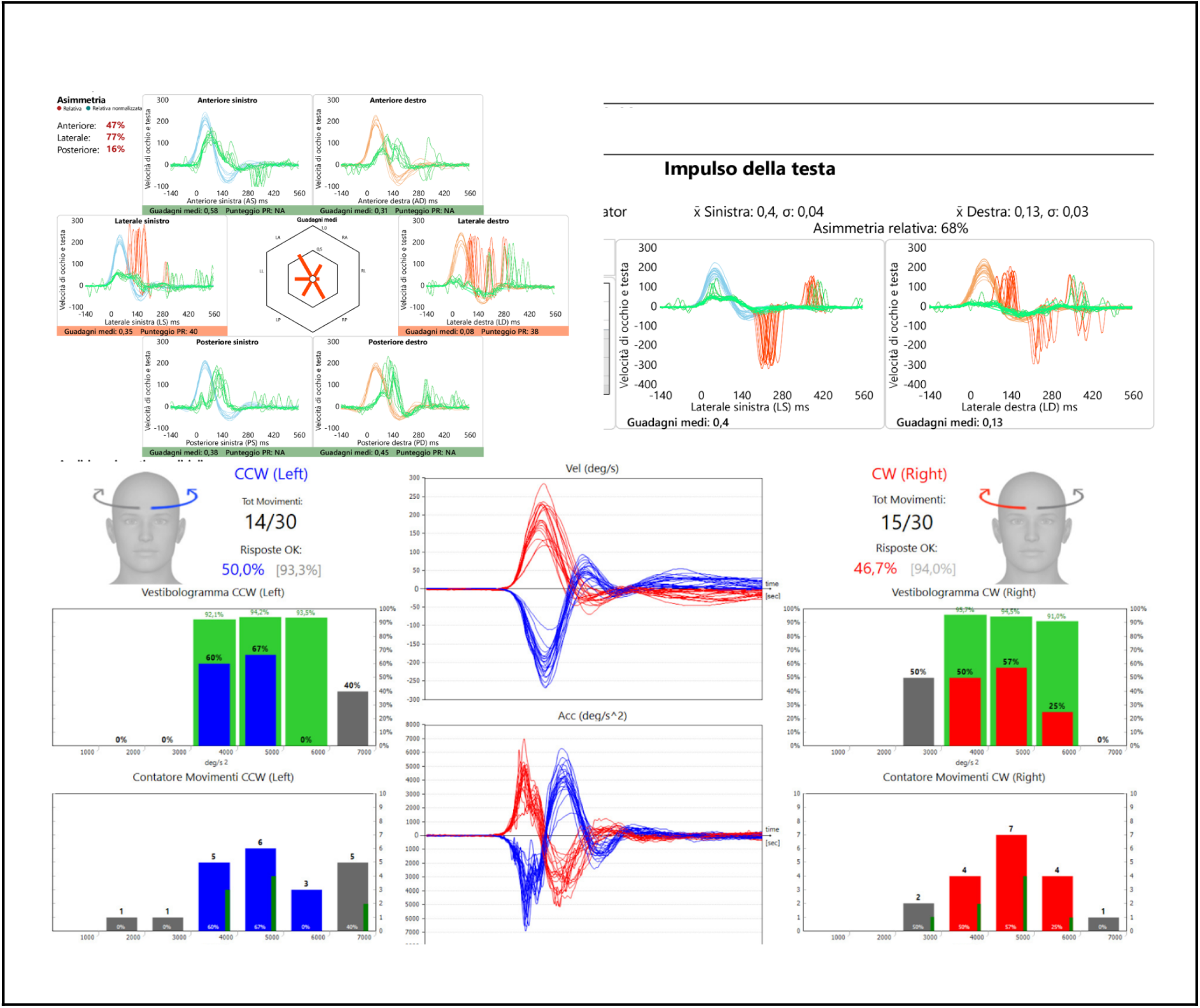
Results from the separate testing protocol. vHIT in the HIMP paradigm demonstrated a bilateral vestibular deficit. SHIMP responses showed reduced gain with the presence of anticompensatory saccades bilaterally. fHIT performance was below normal values on both sides.

**Figure 8:**
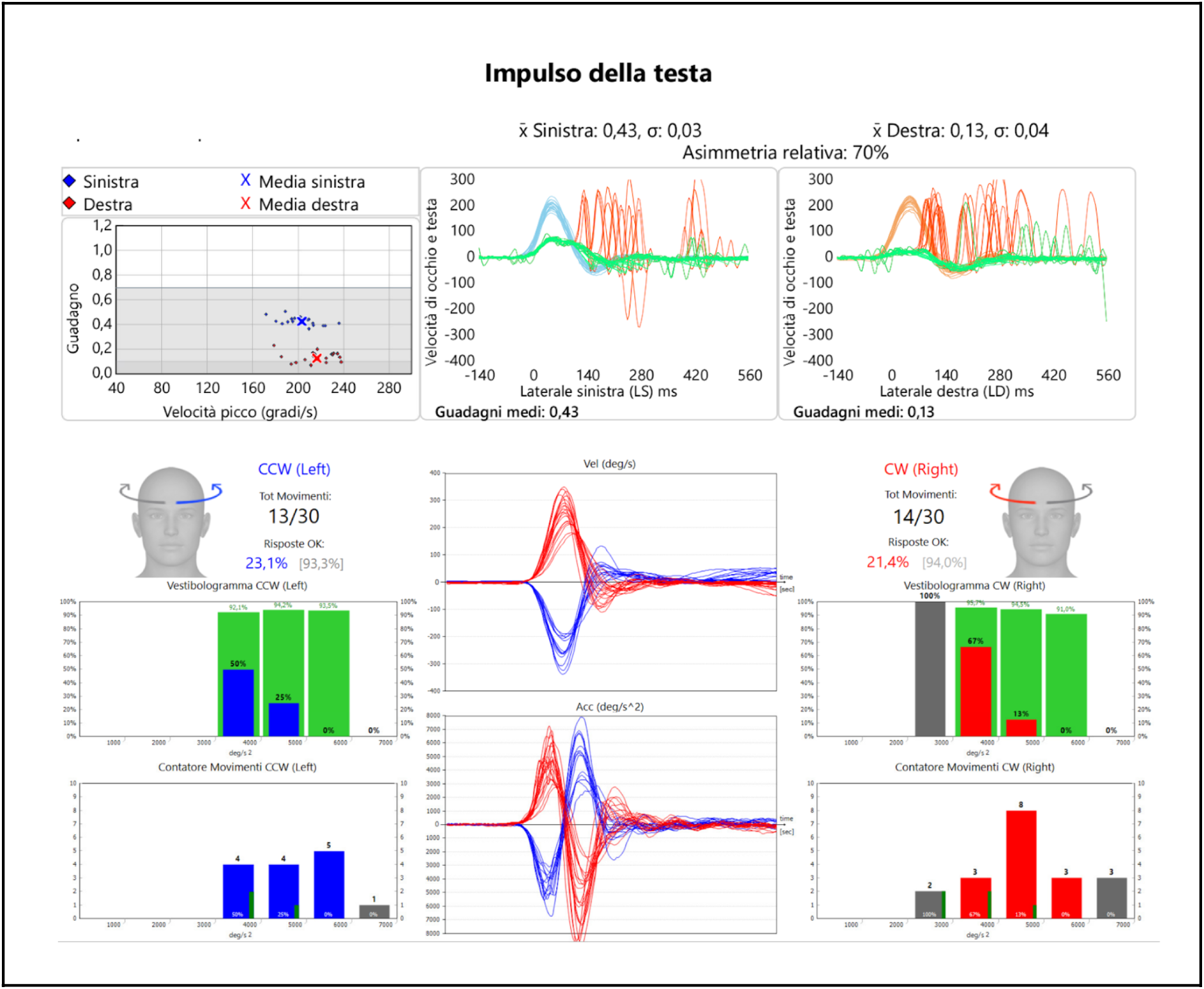
Combined testing protocol. Upper panel: SHIMP responses showing absence of anticompensatory saccadic on both sides. Lower panel: fHIT results demonstrating a marked reduction in the percentage of correct responses during booth head rotations.

Under the separate protocol, the patient compensated for bilateral vestibular areflexia almost exclusively through a highly effective saccadic substitution strategy, capable of largely replacing the slow-phase component, which was severely impaired on the left side and absent on the right.

However, during the simultaneous protocol, the dual-task condition resulted in complete suppression of SHIMP saccades, disrupting a visuo-vestibular interaction that appeared functionally optimal under separate testing conditions.

At the same time, bilateral deterioration of optotype recognition performance was observed.

Overall, these findings suggest increased vulnerability of visuo-vestibular integration under dual-task conditions, consistent with bilateral vestibulopathy and with reduced system capacity to maintain functional stability under additional cognitive load.

### 3.3 Experiment 3

#### 3.3.1 Sample

Ten chronic-phase patients (5 males and 5 females, aged 28–73 years) with previous vestibular insults of different etiologies were enrolled.

#### 3.3.2 Clinical distribution

- 6 unilateral vestibular deficits: 3 right superior vestibular nerve branch AVD, 1 right complete AVD, 2 left superior vestibular nerve branch AVD ( See Table Ca)
- 4 bilateral vestibular deficits (See Table Cb)

#### 3.3.3 Recorded parameters

- VOR gain of the lateral semicircular canals using vHIT in HIMP mode
- Mean latency of HIMP catch-up saccades for both lateral canals
- Clustering of HIMP saccades expressed as the PR index for both lateral canals

These parameters were subsequently recorded again in the same participants using the combined protocol (vHIT-HIMP associated with fHIT). After a few minutes, vHIT in HIMP mode was repeated once more under separate protocol conditions.

**Table Ca.**
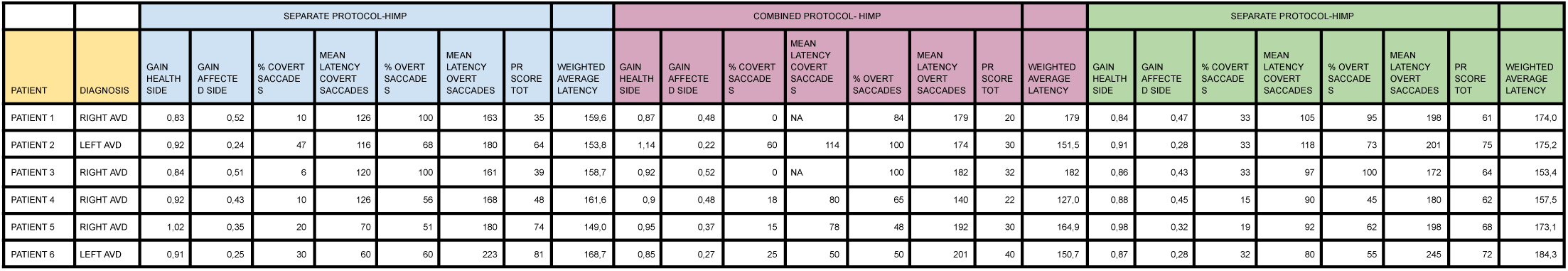
: Results from the third experiment in patients with unilateral vestibular deficits. Parameters referring to the pathological lateral semicircular canal are reported, including healthy-and affected-side lateral canal gains, percentage of overt and covert saccades, mean saccadic latency, PR score, and weighted mean values. Data were collected using the initial separate protocol, the combined protocol, and the final separate protocol, performed approximately 10 minutes apart.

**Table Cb:**
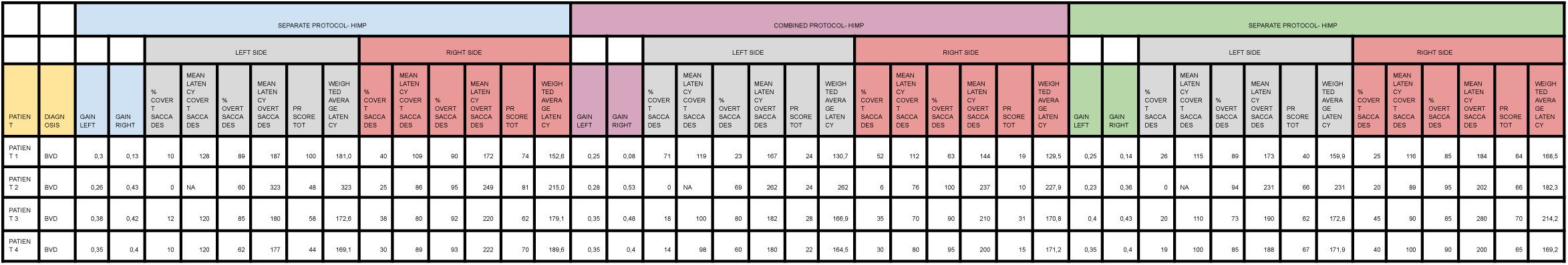
Results from the third experiment in patients with bilateral vestibular deficits. Parameters referring to both affected lateral semicircular canals are reported, including lateral canal gains, percentage of overt and covert saccades, mean saccadic latency, PR score, and weighted mean values. Data were collected using the initial separate protocol, the combined protocol, and the final separate protocol, performed approximately 10 minutes apart.

The objective of this experiment was to evaluate possible variations in mean catch-up saccadic latency and in their degree of clustering, expressed by the PR index. No statistically significant changes in VOR gains were observed either on the healthy or affected side in the unilateral vestibular loss group, nor in the bilateral vestibular deficit group.

In 7 out of 10 subjects, a clear improvement in saccadic clustering was observed during the combined protocol, with a marked reduction in the PR index. This parameter subsequently returned to baseline values during the third vHIT recording in HIMP mode, once the cognitive load associated with the Landolt C recognition task had been removed.

Statistical analysis of weighted mean latency values of catch-up saccades showed no statistically significant differences across the three experimental conditions (initial separate protocol, combined protocol, final separate protocol), neither in patients with unilateral vestibular deficit (p-value= 0.951; p-value= 0.087; p-value= 0.337) nor in patients with bilateral vestibular deficit (p-value= 0.058; p-value= 0.336; p-value= 0.624).

Conversely, changes in saccadic temporal organization, evaluated through the PR index, were statistically significant both in unilateral deficits (p-value= 0.0056; p-value= 0.156; p-value< 0.001) and bilateral deficits (p-value= 0.00055; p-value= 0.630; p-value< 0.001), maintaining significance after Bonferroni correction for multiple comparisons.

In particular, the absence of statistically significant differences between the two separate protocol conditions (pre- and post-combined testing), together with the significant differences observed between each of these conditions and the combined protocol, indicates that the reorganization of saccadic temporal structure observed during the combined protocol was transient and most likely related to the cognitive load associated with the Landolt C recognition task.

Overall, these findings indicate that the introduction of the cognitive task associated with the combined protocol produced a significant temporal reorganization of catch-up saccades, as reflected by the reduction in PR index, without significant modifications in mean saccadic latency, suggesting a task-dependent modulation of compensatory saccadic strategy.

Representative examples are reported below.

**Figure 9:**
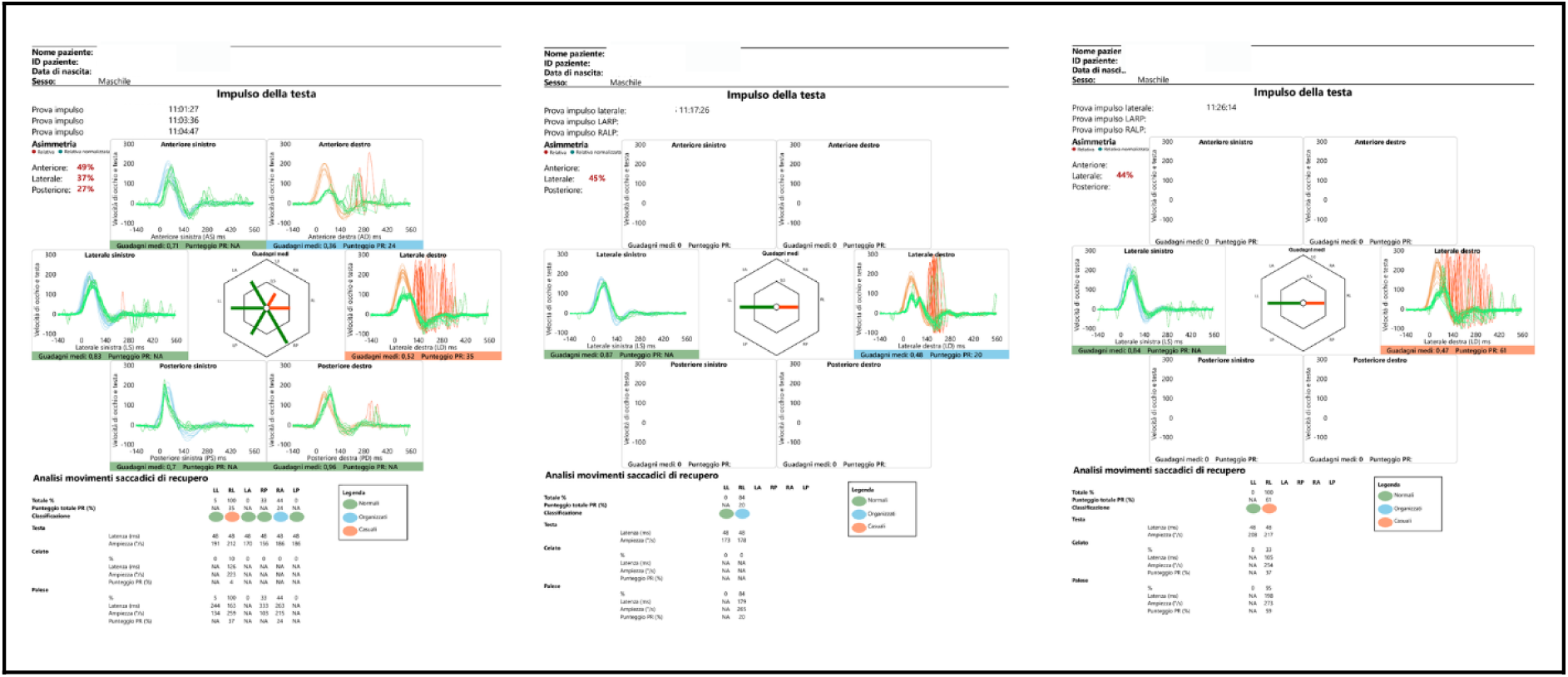
Three vHIT recordings in the HIMP paradigm obtained during the same session in a patient with right superior vestibular nerve dysfunction. Recordings were performed approximately 10 minutes apart and are shown from left to right as follows: initial separate protocol, combined protocol, and final separate protocol. A clear temporal clustering of corrective saccades, reflected by improvement in the PR score, is observed during the combined protocol, followed by an almost complete return to baseline parameters in the final separate protocol.

Three vHIT recordings in HIMP mode were performed at approximately 10-minute intervals.

The first represented the baseline assessment, the second was performed simultaneously with fHIT (combined protocol), and the third was performed approximately 10 minutes after completion of the combined protocol.

During vHIT execution associated with the cognitive recognition task (Landolt C), increased clustering of catch-up saccades was observed, resulting in improvement of the PR index. Specifically, the PR index decreased from 35 in the baseline condition to 20 during the combined protocol and subsequently increased again to 61 during the third vHIT performed without the cognitive task.

**Figure 10:**
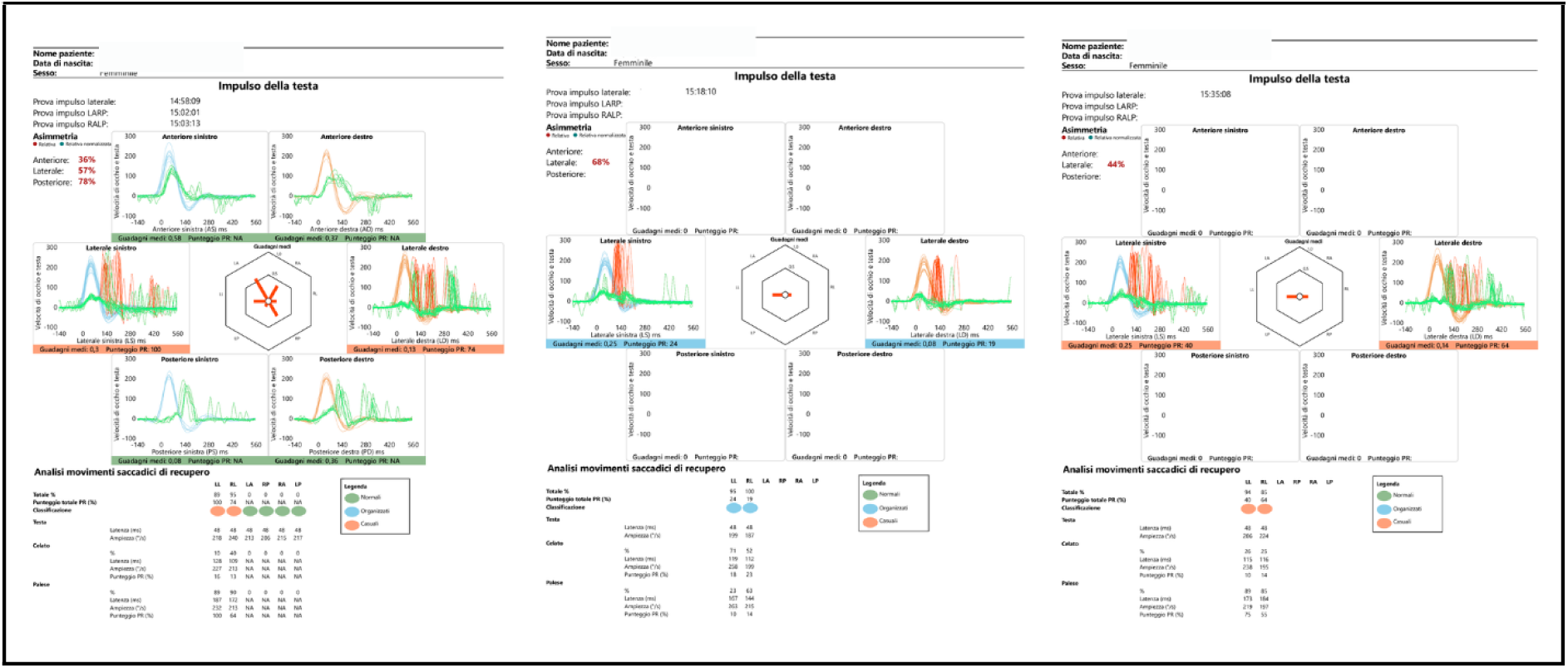
Three vHIT recordings in the HIMP paradigm obtained during the same session in a patient with bilateral vestibular deficit. Recordings were performed approximately 10 minutes apart and are shown from left to right as follows: initial separate protocol, combined protocol, and final separate protocol. A marked temporal clustering of corrective saccades, reflected by improvement in the PR score during the combined protocol, was observed, followed by an almost complete return to baseline values in the final separate protocol.

In a representative patient with bilateral vestibular deficit, weighted mean saccadic latency showed a reduction during the combined protocol compared with the initial HIMP condition, followed by a partial return toward baseline values in the final separate protocol. Specifically, on the left side latency decreased from 181 ms to 130.7 ms during the combined protocol and increased again to 159.9 ms in the final HIMP condition, while PR values markedly improved from 100 to 24 and subsequently partially returned to 40. On the right side latency decreased from 323 ms to 262 ms and then to 231 ms, while PR values changed from 48 to 24 and then to 66.

This pattern supports the presence of a task-dependent improvement in saccadic temporal organization during the combined protocol.

## 4. Discussion

### 4.1 Role of cognitive functions in visuo-vestibular integration

Over the years, vestibular science has progressively benefited from the introduction of numerous technological tools that have consolidated its full integration within the field of neuroscience. Despite the growing body of evidence documenting the significant role of cognitive functions in coordinating and modulating the metrics governing the interaction among different oculomotor reflexes, these tools are still frequently used within an interpretative framework primarily oriented toward quantifying peripheral damage. However, the functional organization of the vestibulo-ocular reflex (VOR) cannot be considered solely as the expression of a low-latency peripheral-brainstem reflex circuit. Rather, it should be interpreted as the result of a dynamic integration with cortical and subcortical oculomotor control systems involved in attentional processes, working memory, and motor prediction mechanisms.

Within this perspective, the SHIMP protocol is commonly used in clinical practice to more sensitively detect deficits of the lateral semicircular canal that, in HIMP mode, may appear less evident due to the presence of compensatory saccadic intrusions. However, the results of the present study suggest that some SHIMP metrics, and in particular the degree of temporal dispersion of anticompensatory saccades, may reflect not only the efficiency of the peripheral vestibular system, but also the level of functional integration between the VOR and extralabyrinthine oculomotor systems responsible for saccadic planning, attentional control, and anticipatory representation of the stimulus.

### 4.2 Effects of cognitive load on eye-head dynamics

The present study explored, using different approaches and, to our knowledge for the first time through an experimental reinterpretation of tools commonly employed in the clinical assessment of balance disorders, the effects of cognitive processes involved in symbolic recognition on visuo-vestibular interaction in healthy subjects and in patients with residual deficits following peripheral vestibular insults of different etiologies. Specifically, working memory and visual attention were intentionally recruited for different functional purposes: in the first two experiments under interference conditions with voluntary saccadic programming, and in the third experiment under facilitation conditions supporting it.

In healthy subjects, the first experiment showed a significant increase in mean SHIMP saccade latency (+85%) during the simultaneous presentation of a complex visual task (Landolt C recognition), without significant changes in the PR index or saccadic amplitude. This finding indicates that visuo-vestibulo-attentional resources were selectively allocated to the visual task at the expense of saccadic generation speed, while preserving the temporal organization of movements.

In patients with chronic vestibular deficit (second experiment), visuo-vestibular interaction appeared more fragile. Half of the subjects maintained SHIMP saccades during the separate protocol, but the introduction of a dual cognitive task sufficient to capture attentional resources produced significant alterations: increased mean saccade latency, temporal disorganization (PR index), disappearance of SHIMP saccades in approximately half of the patients, and reduced optotype recognition performance, particularly on the affected side. In these subjects, a DHI score ≥ 40 was associated with the inability to maintain visuo-vestibular interaction during the dual-task condition, whereas better compensated patients showed greater stability of visuo-vestibular dynamics while accepting a reduction in visual performance. These findings suggest that some apparently favorable oculomotor metrics may not fully reflect residual functional difficulties emerging under more ecologically complex conditions.

The third experiment demonstrated that the targeted integration of working memory and visual attention during the HIMP protocol with a functional task enhances compensatory saccade performance, with a significant reduction in the PR index without changes in mean latency. This reversible effect upon returning to the separate protocol suggests that the oculomotor system adopts flexible compensatory strategies by temporally regrouping saccades to optimize visuo-vestibular integration and preserve visual performance.

From a prognostic perspective, these findings support the development of instrumental assessment protocols capable of more sensitively and specifically monitoring correlations between oculomotor parameters and patient-reported symptoms during chronic phases. In particular, such protocols should more faithfully reflect the complexity of the integrative demands imposed on the visuo-vestibular system during natural head and body movements, in which the interaction among oculomotor reflexes, attentional processes, and working memory represents an essential functional component.

Considering that the PR index represents an indicator of the degree of saccadic clustering, it may be interpreted as one of the earliest expressions of central vestibular compensation, preceding the reduction of saccadic latency and the progressive refinement of their timing relative to the residual deficient slow phase, ultimately leading to the consolidation of automatic, no longer voluntary, responses. Overall, these results suggest the potential rehabilitative value of protocols integrating targeted cognitive activation components, such as those employed in approaches like VGYM, to support functional reorganization of visuo-vestibular interaction.

### 4.3 Neuropsychological models and allocation of attentional resources

Difficulties associated with the simultaneous execution of two or more tasks have led to the development of several neuropsychological models of information processing, including central capacity sharing theory, bottleneck theory, and multiple-resource models. These theories should not be interpreted as alternative explanations, but rather as complementary perspectives describing the same process of limited attentional resource allocation.

Central capacity sharing theory postulates that attentional resources are limited and that the simultaneous execution of two tasks inevitably results in performance deterioration in at least one of them, particularly when the temporal interval between stimuli decreases (37). This theory assumes that, under dual-task demands, attention can be voluntarily allocated to only one specific performance at a time, even when both tasks are highly practiced and largely automatic. This model appears consistent with the results of the first experiment in healthy subjects, in whom the introduction of the optotype recognition task during head rotation produced an increase in SHIMP saccade latency, suggesting functional competition for attentional resource allocation.

Bottleneck theory, on the other hand, proposes that when two tasks involve largely overlapping neural circuits, as in the case of saccadic programming, attentional processing, and working memory, the processing of the second task is temporarily delayed until completion of the first (38). This interpretation is compatible with the increase in mean SHIMP saccade latency observed during the joint protocol in the first experiment, likely attributable to the need to complete symbolic recognition processing before subsequent saccadic response programming.

Interestingly, in the second experiment involving patients with vestibular disorders, the introduction of a dual task revealed specific effects on temporal processing. During the combined protocol, subjects with the worst DHI scores showed disruption of the hierarchical temporal sequence predicted by bottleneck theory: engagement of processing resources in optotype recognition prevented timely release of the resources required to generate efficient anticompensatory saccades, resulting in the disappearance of SHIMP saccades within the temporal window detected by the vHIT software.

Finally, multiple-resource models (39,40) suggest that processing may require the simultaneous engagement of different types of cognitive resources, providing an additional interpretative framework for the observed phenomena.

Taken together, these models support the hypothesis that partial overlap between cortical circuits involved in attentional processes, working memory, and oculomotor control represents a key factor in understanding difficulties in cognitive resource allocation contributing to symptoms reported by many vestibular patients, particularly in dynamic conditions requiring simultaneous integration of multiple sensorimotor tasks. Some researchers suggest that delays may occur only at the response-selection stage, whereas others propose that processing delays may occur at any stage (37, 38).

### 4.4 Ecological and clinical implications of dual-task conditions

Conversely, if two tasks do not share common neural resources, interference between them is unlikely. For example, walking while performing a cognitive task may not significantly alter gait, whereas a second motor task relying on the same resources as walking may generate interference. Based on this principle, we believe that rehabilitation of eye–head dynamics following vestibular insult should include exercises that go beyond simple head rotation while maintaining gaze on a target and should incorporate a contextual cognitive task, such as symbolic image recognition. This approach exploits shared cognitive resources between eye movements and visual processing, stimulating the central nervous system to optimize both performances. In the third experiment, using the combined protocol associating HIMP mode with a functional task, we demonstrated that integration of visual attention and working memory enhances compensatory saccade performance, with immediate improvement in saccadic dispersion as indicated by the PR index.

The effects of dual-task conditions on gait have been widely studied in several populations, including healthy young and older adults as well as patients with neurological disorders (e.g., stroke or brain injury). Most studies in healthy adults report that performing a second task influences gait: dual-task conditions tend to reduce performance in the secondary task and walking speed while leaving other parameters, such as gait variability, unchanged (41).

This pattern of interference in healthy subjects shows similarities with the findings of our first experiment, where worsening of mean SHIMP saccade latency was observed with near preservation of PR index values. Similarly to gait, eye-head dynamics also show a non-zero dual-task cost. Simultaneous execution of two attentional tasks not only produces competition for available cognitive resources but also requires the brain to prioritize between competing demands, highlighting the complexity of neural resource management during multisensory and motor activities.

Two brain regions are commonly involved in defining priorities during simultaneous task execution: the prefrontal cortex (PFC) and the anterior cingulate cortex (ACC). Activation of both has been widely documented in dual-task studies (42,43). Williams proposed that the meaning and relevance of simultaneous information are determined by a fundamental motivation aimed at minimizing danger and maximizing reward (44). This, together with the temporal hierarchy proposed by bottleneck theory, may explain why in the first two experiments both healthy subjects and patients with vestibular insults tended to prioritize optotype recognition over SHIMP saccade efficiency despite explicit instructions to prioritize the saccadic task. Recognition of a continuously changing environment likely represents a primary requirement for maintaining full environmental awareness and generating appropriate behavioral responses.

In patients with vestibular insults observed during the second experiment, marked difficulties in attentional resource allocation emerged. These resources appeared to be “captured” by the attempt to recognize the optotype during head rotation, generating disorder in visuo-vestibular integration. In walking environments rich in interpretative stimuli, this tendency may favor the emergence of symptoms and gait alterations often interpreted as visual dependence, similarly to what has been described in migraine patients. However, a fundamental difference between these two conditions likely exists and is also reflected in rehabilitation behavior. For example, older adults at high risk of falls appear to prioritize planning future actions over accurate execution of ongoing movements, a strategy that may increase fall risk (45). The authors attribute this behavior to anxiety-related processes and the need to gain additional time to analyze the environment before regulating movement. These difficulties resemble those observed in many patients with vestibular insults.

Thus, the phenomenon commonly defined as visual dependence in some vestibular patients may reflect an acquired strategy aimed at anticipating environmental visual information processing as much as possible. This anticipation leads to redistribution of attentional resources that may sometimes be disadvantageously withdrawn from the ongoing motor act, resulting in a disturbance of vestibular processing. Our data suggest that this behavior may be modulated through targeted dual-task interventions designed to enhance attentional resource allocation capacity in these subjects, thereby improving visuo-vestibular integration and motor efficiency.

### 4.5 Rehabilitation implications

The results of the three experiments confirm the central role of saccadic movements in compensation for vestibular deficits and the influence of cognitive processes on oculomotor kinematics. The PR index, an indicator of saccadic clustering, emerges as an early marker of central compensation, preceding latency reduction and refinement of saccadic timing. Integration of targeted cognitive activation components, as implemented in the VGYM protocol, may therefore facilitate functional reorganization of visuo-vestibular dynamics.

Overall, the combined use of SHIMP, HIMP, and fHIT allows detection of subtle interactions between cognitive functions and oculomotor control, providing diagnostic and rehabilitative tools that are more sensitive and closer to the ecological conditions of everyday life.

## 5. Conclusions

Taken together, the results of the present study confirm that visuo-vestibular integration cannot be interpreted exclusively as the expression of peripheral-brainstem reflex mechanisms, but should instead be considered the result of a complex dynamic interaction among oculomotor systems, attentional processes, and working memory. In particular, the findings demonstrate that the kinematics of SHIMP saccades and compensatory catch-up saccades represent not only indicators of residual vestibular function, but also reflect the efficiency of cognitive processes involved in movement planning and attentional resource allocation.

The introduction of symbolic-interpretative cognitive tasks significantly modulated both saccadic latency and temporal dispersion in healthy subjects as well as in patients with residual peripheral vestibular deficits. In healthy subjects, dual-task conditions primarily resulted in increased mean saccadic latency without altering the temporal organization of responses, suggesting selective reallocation of attentional resources. In contrast, in patients with chronic vestibular disorders, cognitive interference revealed greater fragility of visuo-vestibular integration, with temporal disorganization of anticompensatory saccades and, in the most symptomatic cases, disappearance of SHIMP saccades under conditions of increased attentional load. These findings indicate that some oculomotor metrics that appear favorable under standard testing conditions may not adequately reflect functional difficulties emerging in more ecologically complex contexts.

At the same time, the targeted integration of working memory and attention during the execution of oculomotor tasks showed a facilitating effect on the clustering of compensatory catch-up saccades, as documented by the reduction of the PR index. This observation suggests that targeted cognitive activation may represent a relevant modulatory factor in functional reorganization processes and, precisely because it is modifiable, may be effectively exploited for rehabilitative purposes within vestibular rehabilitation programs.

From a clinical and prognostic perspective, these findings support the usefulness of instrumental protocols incorporating dual-task conditions in the assessment of eye-head dynamics, in order to identify early alterations of visuo-vestibular integration that may not be detectable using standard testing paradigms. In particular, the saccadic clustering index (PR) emerges as a potential early biomarker of central compensation processes within the HIMP paradigm and as a parameter sensitive to variations in attentional resource allocation.

Finally, the present findings suggest important rehabilitative implications: the systematic integration of contextual cognitive tasks into gaze-stabilization exercises may promote more effective enhancement of central compensatory mechanisms, making vestibular rehabilitation more closely aligned with the multisensory demands of everyday life. From this perspective, rehabilitation protocols combining head rotations with symbolic recognition or visually interpretative tasks may represent a promising evolution of traditional approaches, contributing to improved efficiency of visuo-vestibular interaction and better functional recovery outcomes in patients with peripheral vestibular deficits.

## Abbreviations

DHI: Dizziness Handicap inventory
vHIT: video Head Impulse Test
SHIMP: Suppression Head Impulse Protocol
HIMP: Head Impulse Protocol
fHIT: functional Head Impulse Test
VOR: vestibulo-ocular reflex
MMSE: Mini-Mental State Examination
HSC: horizontal semicircular canals
SSC: superior semicircular canals
PSC: posterior semicircular canals
PR: Perez and Rey
AVD: acute vestibular deficit
BVD: bilateral vestibular deficit
VGYM: VOR gym
PFC: prefrontal cortex
ACC: anterior cingulate cortex
NA: not applicable
PPPD: persistent postural-perceptual dizziness

## Data Availability

All data generated or analyzed during this study are included in this published article. Supplementary information is available on request from the corresponding author.

## Acknowledgments

We would like to thank the participants who took part in this study for their time and cooperation.

## Author contributions

P.M.: original idea, general coordination, review and conducted most data acquisition. A.G.T: contributed to the manuscript writing and editing, data analysis and table editing and figures. A.C, S.M, L.M. e M.M: contributed to the final editing. All authors made substantive intellectual contributions to the manuscript and revised it. All authors have read and agreed to the published version of the manuscript.

## Statements and declarations

### Ethical considerations

This retrospective study was approved by the AVEN Ethics Committee (approval number for the promoter institution: 792/2021/OSS/AUSLRE). Participants included in the analysis were recruited between May 2020 and April 2021 and had provided written informed consent prior to enrolment in the study, including consent for the use of their data for research purposes. This research was conducted ethically in accordance with the World Medical Association Declaration of Helsinki.

### Consent to participate

Written informed consent has been obtained from the patients to participate in this paper.

### Consent for publication

Written informed consent has been obtained from the patients to publish this paper.

